# Evaluation of SARS-CoV-2 Antibody Point of Care Devices in the Laboratory and Clinical Setting

**DOI:** 10.1101/2021.12.16.21267703

**Authors:** Kirsty McCance, Helen Wise, Jennifer Simpson, Becky Bachelor, Harriet Hale, Lindsay McDonald, Azul Zorzoli, Elizabeth Furrie, Charu Chopra, Frauke Muecksch, Theodora Hatziioannou, Paul D. Bieniasz, Kate Templeton, Sara Jenks

## Abstract

SARS-CoV-2 Antibody tests have been marketed to diagnose previous SARS-CoV-2 infection and as a test of immune status. There is a lack of evidence on the performance and clinical utility of these tests. We aimed to carry out an evaluation of 14 point of care (POC) SARS-CoV-2 antibody tests.

Serum from participants with previous RT-PCR (Real-Time Polymerase chain reaction) confirmed SARS-CoV-2 infection and pre-pandemic controls were used to determine specificity and sensitivity of each POC device. Changes in sensitivity with increasing time from infection were determined on a cohort of participants. Corresponding neutralising antibody status was measured to establish whether the detection of antibodies by the POC device correlated with immune status. Paired capillary and serum samples were collected to ascertain whether POC devices performed comparably on capillary samples.

Sensitivity and specificity varied between the POC devices and in general did not meet the manufacturers reported performance characteristics signifying the importance of independent evaluation of these tests. The sensitivity peaked at >20 days following symptoms onset however sensitivity of 3 POC devices evaluated at extended time points showed that sensitivity declined with time and this was particularly marked at >140 days post infection onset. This is relevant if the tests are to be used for sero-prevelence studies. Neutralising antibody data showed positive antibody results on POC devices did not necessarily confer high neutralising antibody titres and these POC devices cannot be used to determine immune status to the SARS-CoV-2 virus. Comparison of paired serum and capillary results showed that there was a decline in sensitivity using capillary blood. This has implications in the utility of the test as they are designed to be used on capillary blood by the general population.

## Introduction

The emergence of SARS-CoV-2 has triggered a race by laboratories and manufacturers to bring commercial diagnostic tests to market. The unusually rapid pace of this development risks potential compromises on quality in the absence of rigorous independent evaluation and validation. It is therefore essential to ensure adequate performance of tests for population wide or individual use to prevent roll out of tests which add no, or at best, minimal clinical value to individual patients or the wider population.

The current most widely used diagnostic test for SARS-CoV-2 is based on real-time polymerase chain reaction (RT-PCR) amplification of viral RNA from an upper respiratory tract sample (1,2). However, due to a limited time window of active infection, capacity constraints and restriction of access to symptomatic patients, cases determined using this method underestimate the true burden of infection. In contrast, serological assays test for previous infection and are therefore a key additional tool for monitoring prevalence of infection within the population. Antibody tests can also be used as an aid in diagnosis where COVID-19 is suspected clinically but the PCR time window has passed (3) and significant interest exists in the potential for use of these tests at an individual level to provide an indication of immune status and act as an ‘immunity passport’. For countries where vaccine availability is limited pre-screening the population with antibody tests to identify individuals who may either not require vaccination or be suitable for a reduced vaccine dosing regimen may help optimise the use of limited vaccine resources.

Of particular interest is the use of monoclonal antibody treatment in hospitalised patients with SARS-CoV-2. Ronapreve, a combination of Casirivimab and Imdevimab, is a monoclonal antibody treatment directed at the Spike protein receptor binding domain on SARS-CoV-2.

This has been shown to significantly reduce 28 day mortality in seronegative hospitalised patients (4).Therefore a well performing antibody tests has the potential to identify patients who would benefit from this treatment.

A vast number of commercially available immunoassays have been developed to detect SARS-CoV-2 IgG, IgM, IgA and total antibodies (5). Although the majority of antibody production is directed towards the more abundant N (nucleocapsid) protein, the S (spike) protein contains the receptor-binding domain responsible for host cell attachment and antibodies to the S protein are therefore predicted to be neutralising (6).

In contrast to laboratory-based immunoassays, which require venous sampling and transport to centralised testing sites, lateral flow immunoassays (LFA) offer the potential to allow rapid, cheap, mass population antibody testing on capillary samples in the home environment. This relief on clinical pressure and its use at a population level relies on the LFA working on capillary whole blood samples and the tests offering sufficient ease of use, interpretation and acceptability to the wider population. If LFAs are to be used for population sero-surveillance they will have to be sensitive enough to detect the presence of antibodies in those who only suffered from mild disease or were asymptomatic. Of even greater importance is the test specificity when testing at a population level where the pre-test probability is low. Without adequate specificity the chance of a positive result being a false positive is considerable and this is of particular concern if these tests were to be used as evidence of immune status or ‘immunity passports’. It is currently unclear if the detection of antibodies to the COVID 19 virus by LFA confers immunity to the virus and some manufactures fail to disclose whether the assay is directed towards the N or S protein.

Worldwide, over 200 LFAs have been produced to date (5), for the majority of which only manufacturer reported performance data is available. There is no requirement for external validation to obtain CE marking and the manufacturer’s validation process is often not publically available. The UK Medicines and Healthcare Products Regulatory Agency (MHRA) advise that any SARS-CoV-2 LFA should have sensitivity and specificity > 98% (95 % CI 96-100 %) in samples collected over 20 days post symptom onset irrespective of whether they are performed as a home self test kit or by a trained health care professional (7). Despite claims by many manufactures to have achieved this target (see S1), UK external validations of LFAs have so far failed to reproduce this (8–10).

As part of a Scottish evaluation of SARS-CoV-2 diagnostic tests, we carried out an independent validation of 14 point of care SARS-CoV-2 antibody tests including 13 LFAs. We assessed their specificity and sensitivity against manufacturer’s claims and determined compatibility with MHRA criteria. For tests that performed well against the MHRA criteria, we studied changes in sensitivity with increasing time from infection. To investigate suitability for home use, we evaluated ease of use and interpretation of these tests when performed directly by patients. Neutralising antibody status was determined for samples to establish whether the detection of antibodies by the LFA correlated with immune status.

## Methods

### Point of care (POC) device selection and the evaluation process

Companies approached National Services Scotland (NSS) with their POC device. The manufacturer claims and costing were reviewed and manufacturers who passed these initial checks were invited to send up to 100 POC test kits for initial evaluation. 14 point-of-care devices designed to detect antibodies to the SARS-CoV-2 virus were evaluated. The tests included in the evaluation were AbC-19, Alpha Pharma, Biomerica, Biozek, Fortress, Jiangsu, Lepu, Menarini Healgen, Mologic, Pharmact, Roche, Wuhan Easy Diagnosis, Wuhan Life Origin Biotech (Syzbio) and LumiraDX. 13 of the POC devices were LFAs, consisting of immunochromatography based cassettes, with the LumiraDX assay using a point of care microfluidic immunofluorescence assay. Each device measured IgG and IgM except Abc19 which only measured IgG and LumiraDX which gave an antibody result of unspecified subclass. Target antigens (either S or N) were not always disclosed by the manufacturers. The tests were performed as per the manufacturer’s instructions. Typically, 2.5µL to 10µL of serum or capillary sample was pipetted into the sample well followed by a pre-specified volume of buffer supplied by the manufacturer (see S1). Serum samples were run at room temperature in the laboratory at the Royal Infirmary of Edinburgh and the read time varied from 10 to 20 minutes depending on manufacturer’s instructions. Photographic records of serum results were taken. Capillary samples were run as part of a research clinic. Antibody results were visually read and recorded as positive, weak positive, or negative for IgG and IgM. Positive and weak positive results were deemed an overall positive result.

In the first stage of the process 50 serum samples from RT-PCR confirmed SARS-CoV-2 positive participants and 50 negative controls were run. Initial sensitivity and specificity were calculated and if the test kit performed well (typically defined as an IgG specificity of >98% and sensitivity of >95% at >day 21 post infection) the company was taken forward to the second stage of evaluation and asked to provide further test kits. Further stages of the evaluation process which were carried out for a subset of kits included more stringent specificity testing, determination of batch to batch variation, sensitivity as time from infection increased, comparison of capillary to serum results and evaluation of ease of use by study participants in a research clinic.

### Overview of participants and samples

Serum samples from patients with RT-PCR confirmed SARS-CoV-2 were collected between March and November 2020. Excess serum was stored at the point of discard from hospitalised COVID-19 patients using ethical permissions obtained through NRS BioResource.

Out-patients with previous RT-PCR confirmed SARS-CoV-2 infection were also recruited to two COVID convalescent research studies where participants were invited to donate serial blood samples (SR1407 BioResource study, n=112 participants) and to provide capillary and serum blood samples (COVID-19 Antibody Test Evaluation (CATE) Study, n= 82 participants). For both studies the date of symptom onset and positive RT-PCR test result were recorded. Inclusion criteria were those aged >16 years old with previous RT-PCR confirmed diagnosis of COVID-19. Only 5 study participants were diagnosed with COVID-19 in hospital with the remainder being diagnosed in the community. Due to the limited access to COVID-19 testing at the start of the pandemic when most study recruitment occurred many study participants were health care workers. Exclusion criteria were any frail or shielding individuals. Each participant provided between 1 and 5 serum samples. For participants providing serial samples these were collected between 24 and 216 days post PCR confirmed infection. Capillary samples were collected as part of the CATE study research clinic between September and November 2020. Capillary samples were tested in the clinic setting with paired serum samples subsequently run on the same POC devices in the laboratory to assess concordance on capillary and serum samples. A subset of CATE study participants were asked to carry out one of the POC tests themselves including finger prick testing with the aid of the manufacturers printed instructions. Health care workers intervened if the participant needed support in completing the testing and any intervention was recorded. Afterwards participants were asked to complete a questionnaire on their experience of performing the test. For a subset of other test kits capillary results were interpreted by both the participant (with the aid of a labelled diagram) and the researcher.

Negative controls consisted of venous samples collected prior to December 2019. For more stringent specificity testing samples positive for rheumatoid factor, seasonal coronaviruses, other respiratory pathogens, CMV (cytomegalovirus) or EBV (Epstein-Barr Virus) were used as negative controls along with samples from the national antenatal screening programme and BTS (The British transfusion service).

Wherever possible the same panel of samples were run on each POC device but due to limitations in sample volume not all devices were assessed with the same panel of samples. However, a similar variety of samples types were used to test each kit.

### Neutralisation assay

Neutralising antibody assays were performed on serum samples using a pseudotyped chimeric virus, expressing the SARS-CoV-2 spike protein, as described in (11).

### Statistical analysis

The primary outcome was the sensitivity and specificity of each POC test. For sensitivity, tests were compared against PCR-confirmed SARS-CoV-2 infection. Specificity of each POC test was evaluated against pre-pandemic negative samples, with all positives counting as false positives. The analysis included all available data for the relevant outcome and are presented with the corresponding 95% confidence intervals (Clopper Pearson).

For samples where neutralising antibody levels are available positive predictive value (PPV) and negative predictive value (NPV) are calculated at two NT_50_ thresholds; ≥ 50 and ≥ 160.

For the comparison of the performance of each POC test between clinic capillary and serum samples we calculated the sensitivity and 95% confidence interval. The McNemar test was used to assess for significant difference between dependent groups. Agreement between serum and capillary samples was measured using the Cohen’s Kappa. Results of the Kappa were interpreted as previously described (<0, poor agreement; 0.00–0.20, slight agreement; 0.21–0.40, fair agreement; 0.41–0.60, moderate agreement; 0.61–0.80, substantial agreement; and >0.8, almost perfect agreement) (12).

All data were analysed using PRISM version 9 and SPSS, and a p value<0.05 was considered significant.

### Ethics

Ethical approval for access to pre-pandemic stored samples and hospitalised COVID-19 patient samples was obtained through the NRS BioResource (Ref: SR1410). Outpatient COVID-19 convalescent patient samples were obtained through the NRS BioResource (Ref: SR1407) and the COVID-19 Antibody Test Evaluation (CATE) study, with ethical approval from London-Brent Research Ethics Committee (REC ref: 20/HRA/3764 IRAS: 286538

## Results

### Comparison of POC device performance using serum samples

14 POC devices were evaluated for sensitivity and specificity including 13 LFAs and one microfluidic immunofluorescence assay (Lumira DX). In the first stage the sensitivity of the LFA was assessed using a panel of 50 serum samples from RT-PCR positive individuals. The specificity of the LFA was assessed using a panel of 50 serum samples collected prior to December 2019 for routine virological investigations. Adequately performing LFA then progressed to stage 2, where they were evaluated against an expanded panel of serum samples. In particular, a more stringent specificity panel was used, including serum samples taken from individuals with a positive PCR result for the endemic coronaviruses, other respiratory pathogens, and samples with positive rheumatoid factor, CMV or EBV serology.

The results are summarised in **Table 1** and **Fig 1**. For each sample, and each device, the IgG and IgM result was scored as negative, weak positive or positive (the Abc19 LFA only had an IgG window, and the LumiraDX platform only returned an antibody result). For the purposes of the sensitivity and specificity calculations, weak positive and positive results were counted as positive.

**Table 1:**
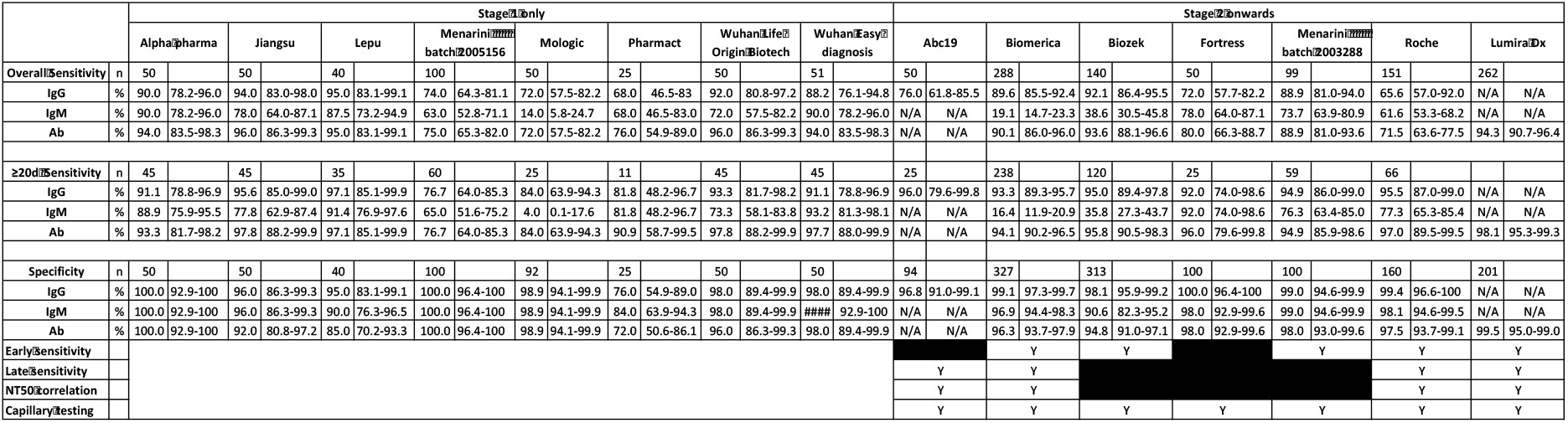
Summary of POC test sensitivity and specificity. For each manufacturer the n, overall sensitivity, sensitivity at ≥ 20 days post symptom onset, and specificity is shown, with 95% confidence intervals (Clopper-Pearson). The LFA are grouped according to which stage of the evaluation they reached. For kits that progressed further than stage 1, the additional analyses performed are indicated (Y, performed). For Abc19, only IgG data was available, and for LumiraDX, only the overall antibody result was available. The results from two separate batches of Menarini kits are also shown.

**Fig 1:**
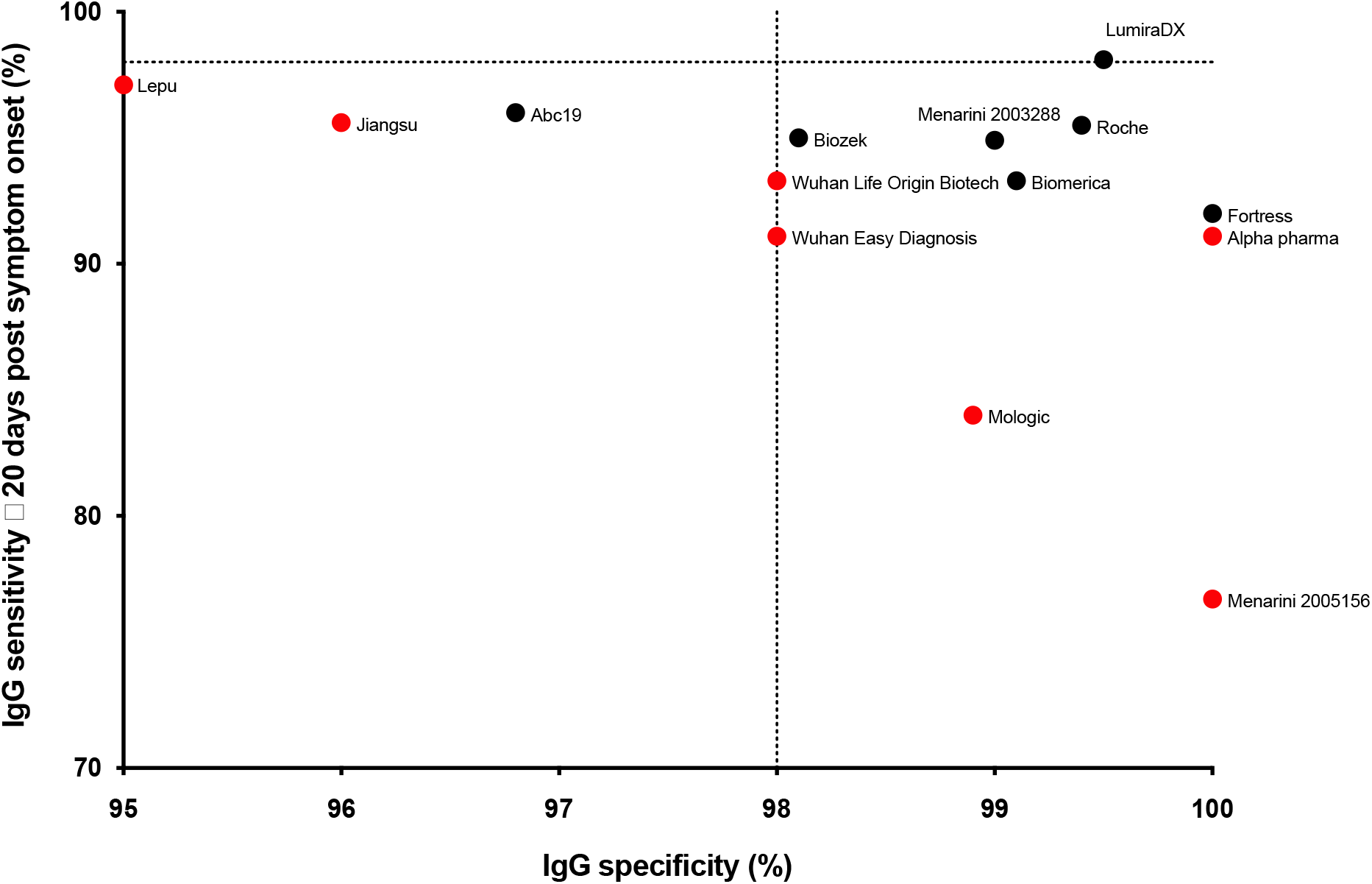
Sensitvity vs specificity at >20 days. Sensitivity against specificity for the kits tested (IgG only, and positive or negative result for the LumiraDx assay). For each kit the ≥ 20 days post symptom onset sensitivity is shown. The MHRA targets of 98% sensitivity and specificity are shown as dotted lines. Pharmact was not included on the graph due to low specificity. The LFA that did not progress beyond this stage are shown in red.

Only the LumiraDX POCT assay reached the MHRA criteria for sensitivity and specificity (> 98% for both), although a number of the LFA (Biomerica, Biozek, Fortress, Menarini and Roche) surpassed the specificity criteria, even when tested against the high stringency specificity serum samples. One empirical finding was that there was extensive variation in the strength of the staining between the kits, which could affect ease of interpretation. Another more worrying observation was that batch-to-batch variation in performance was observed for the Menarini assay with batch 2005156 giving a sensitivity of 76.3% on samples ≥20 days post infection versus batch 2003288 giving a sensitivity of 94.9% on the same samples (n=99).

The Abc-19, Biomerica, Biozek, Fortress, Menarini (batch 2003288) and Roche LFA and the LumiraDX assay underwent further analysis using an expanded panel of RT-PCR positive samples. Initially, sensitivity was examined at proximal time points to infection for the Biomerica, Biozek, Menarini, Roche and LumiraDX LFAs and the results are shown in **Table 2** and **Fig 2**. All the LFA showed an initial increase in sensitivity, peaking at ≥ 21 days post symptom onset, with sensitivities of ≥ 90% obtained for all LFA from this time point onwards. Between 8-20 days post symptom onset, LFA performance was more variable, with Roche showing the lowest sensitivity, and Menarini the highest.

**Table 2.** Sensitivity against time.

| Post symptom onset (days) |  | Biomerica | Biozek | Menarini | Roche | Lumira |
| --- | --- | --- | --- | --- | --- | --- |
| 0-7 | Sensitivity | N/A | N/A | N/A | **11.5** | **26.1** |
|  | 95% CI | N/A | N/A | N/A | 2.4-27.2 | 10.2-45.1 |
|  | n | N/A | N/A | N/A | 26 | 23 |
| 8-20 | Sensitivity | **77.2** | **81.8** | **83.3** | **59.4** | **71.6** |
|  | 95% CI | 64.2-85.9 | 59.7-93.5 | 69.8-91.4 | 46.4-69.7 | 60.0-80.1 |
|  | n | 57 | 22 | 48 | 64 | 74 |
| 21-40 | Sensitivity | **93.0** | **95.6** | **100** | **93.8** | **92.9** |
|  | 95% CI | 83.0-97.6 | 87.6-98.8 | 84.6-100 | 79.2-98.9 | 82.7-97.5 |
|  | n | 57 | 68 | 22 | 32 | 56 |
| 41-60 | Sensitivity | **95.2** | **95.1** | **89.7** | **96.6** | **98.7** |
|  | 95% CI | 88.3-98.4 | 83.1-98.3 | 72.6-97.1 | 82.2-99.8 | 93.1-99.9 |
|  | n | 84 | 49 | 29 | 29 | 78 |

*Sensitivity against time post symptom onset for selected kits (IgG for all LFA, with the exception of LumiraDX, where overall antibody result was used). n and 95% confidence intervals (Clopper Pearson) are shown. N/A, not done.*

**Fig 2.**
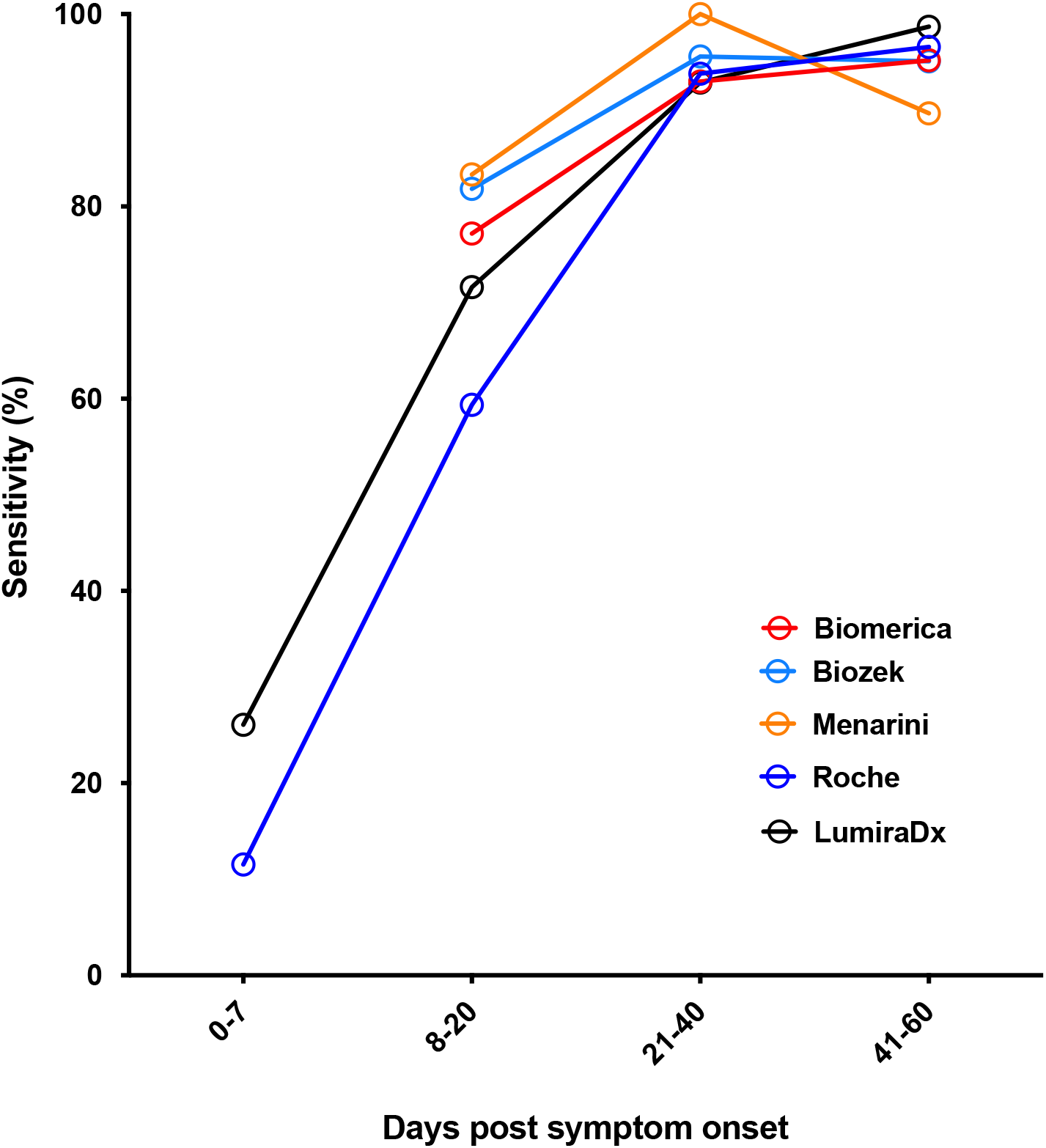
Sensitivity against time. Sensitivity against time post symptom onset for selected kits (IgG for all LFA, with the exception of LumiraDX, where overall antibody result was used).

An equally important question was what the sensitivity of the POCT LFA was at later time points post symptom onset, since it has been shown that antibody levels wane with time following the primary antibody response to SARS-CoV-2 (13–15). Sensitivity was analysed across three time windows from 20 days post symptom onset for the Abc19, Biomerica, Roche and LumiraDX predominantly using samples from the convalescent cohort, and the results are shown in **Table 3** and **Fig 3**.

**Table 3.** Sensitivity against time post symptom onset.

| Post symptom onset (days) |  | Abc19 | Biomerica | Roche | Lumira |
| --- | --- | --- | --- | --- | --- |
| 20-79 | Sensitivity | **80.3** | **91.1** | **98.1** | **98.0** |
|  | 95% CI | 73.0-85.4 | 85.4-94.5 | 94.4-99.5 | 94.4-99.5 |
|  | n | 152 | 157 | 154 | 153 |
| 80-139 | Sensitivity | **81.4** | **88.6** | **93.2** | **97.7** |
|  | 95% CI | 66.6-90.4 | 75.4-95.4 | 81.3-98.12 | 87.7-99.9 |
|  | n | 43 | 44 | 44 | 43 |
| ≥ 140 | Sensitivity | **68.5** | **54.4** | **82.3** | **98.2** |
|  | 95% CI | 56.6-77.4 | 42.8-64.0 | 72.1-88.9 | 90.6-99.9 |
|  | n | 73 | 79 | 79 | 57 |

*Sensitivity against time post symptom onset for selected kits (IgG for all LFA, with the exception of LumiraDX, where overall antibody result was used). n and 95% confidence intervals (Clopper Pearson) are shown. N/A, not done.*

**Fig 3.**
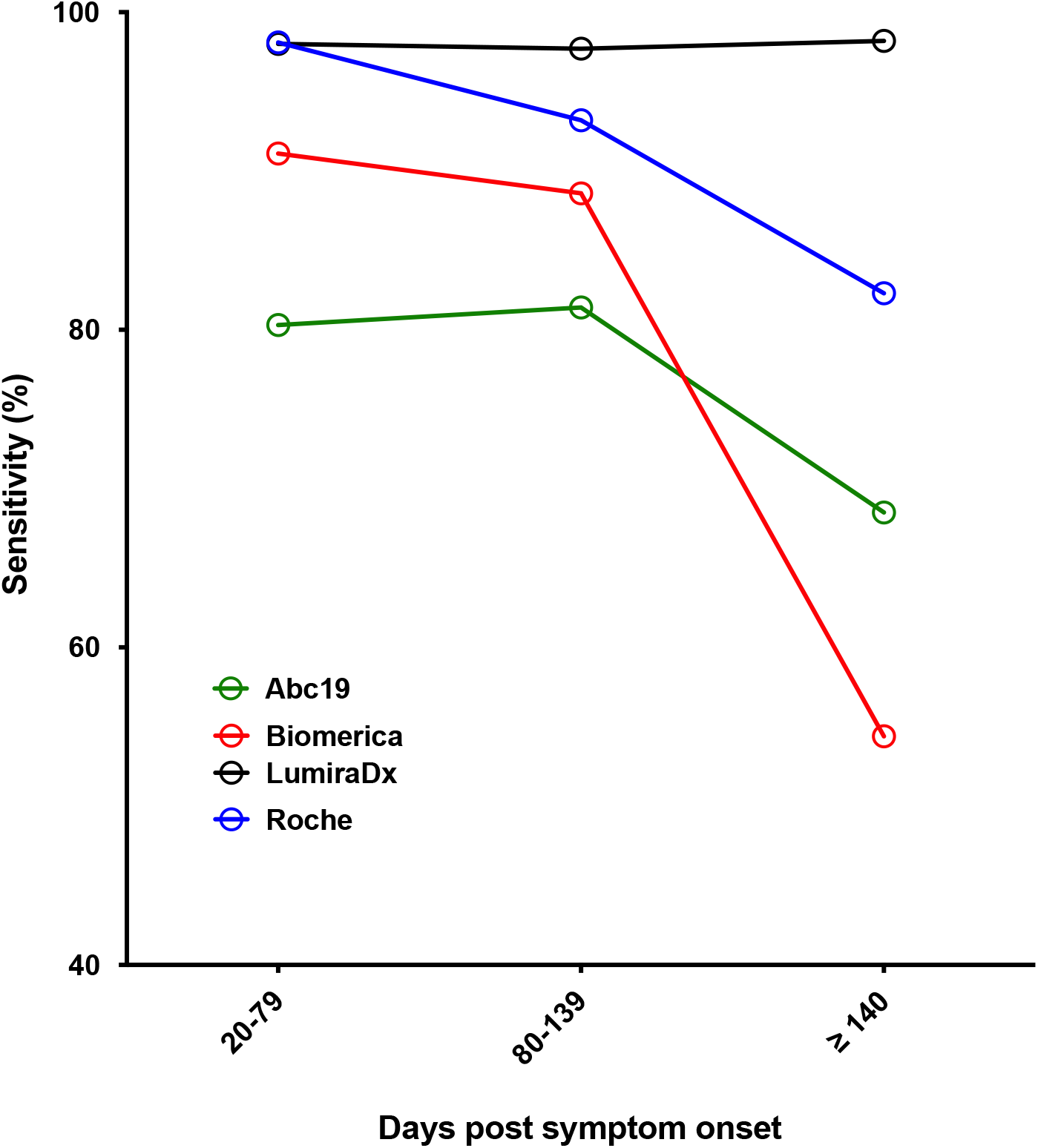
Sensitivity against time post symptom onset. Sensitivity against time post symptom onset for selected kits (IgG for all LFA, with the exception of LumiraDX, where overall antibody result was used).

The LumiraDX assay was the only assay evaluated that maintained its sensitivity at 98% across the time interval studied (up to 224 days post symptom onset). The three LFA showed variable drops in sensitivity with increasing time from symptom onset, with Biomerica dropping from 91% to 54% over the time points studied.

For the convalescent cohort samples, neutralising titres were also available, and the POCT result (for Abc19, Biomerica, Roche and LumiraDX) could be compared with the half maximal neutralising titre (NT_50_). Initially, the NT_50_ result for each serum sample was plotted, dividing each LFA by positive and negative IgG (or for LumiraDX, antibody) result (**Fig 4**).

**Fig 4.**
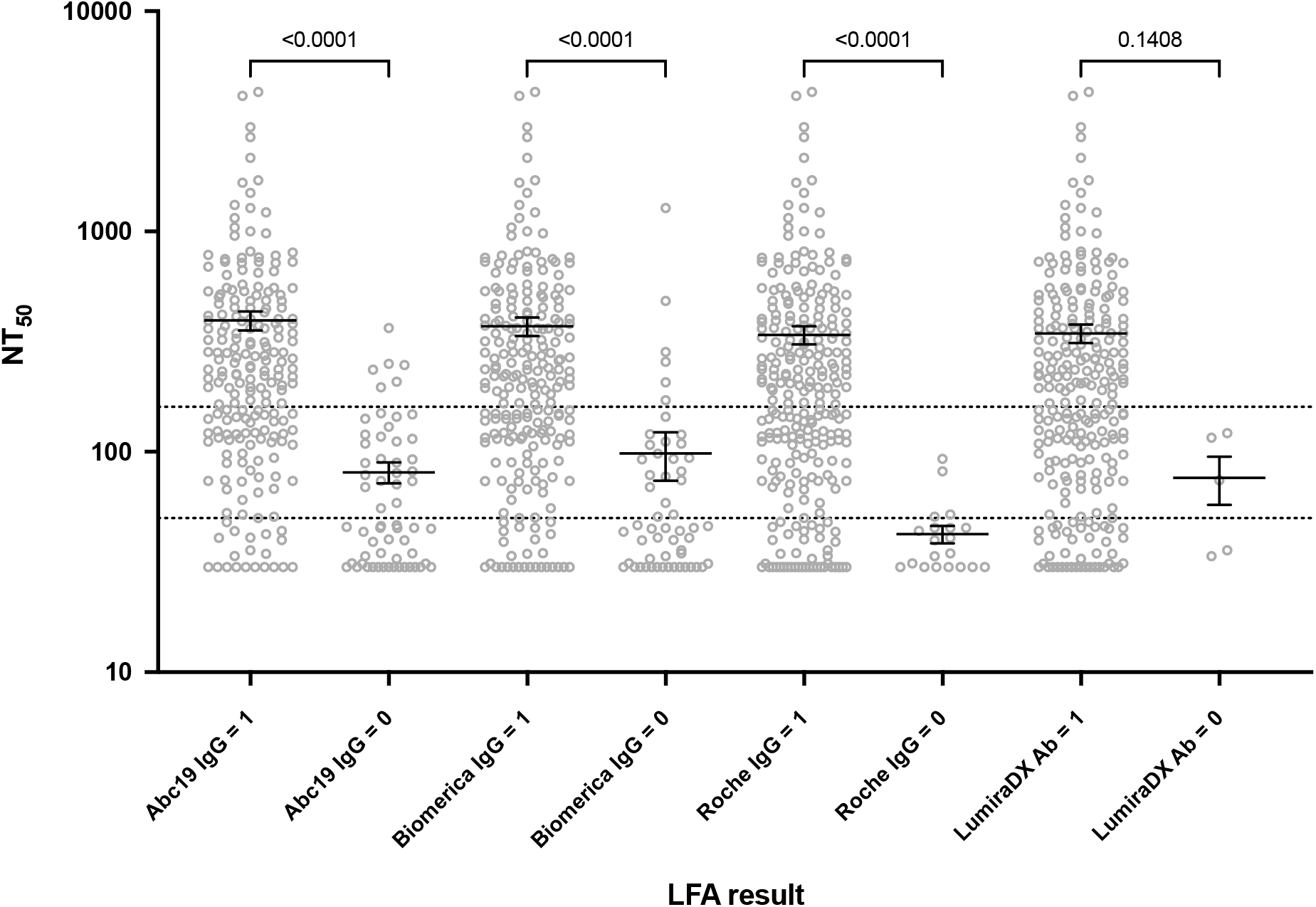
Neutralising antibody titres compared to POC test result. NT_50_ against binary POC test result. The NT_50_ results for the positive and negative LFA result are plotted by kit (the mean and SEM are denoted by black lines). Dotted lines represent the NT_50_ ≥ 50 and ≥ 160 thresholds, and the p values from a Kruskal-Wallis test are shown.

There was a statistically significant difference in NT_50_ level between the positive and negative LFA result for all assays. However, for the LFA positive result, a large range of NT_50_ values was observed. To examine this further, the sensitivity and specificity of the LFA was evaluated at two NT_50_ thresholds; ≥ 50 and ≥ 160 (**Table 4**). These two thresholds were selected based on a detectable NT_50_ (≥ 50) and a threshold which may confer protection from reinfection (≥ 160) (16).

**Table 4.** Performance of POC devices compared to NT_50_ ≥ 50 and ≥ 160.

|  | NT50 ≥ 50 |  |  |  | NT50 ≥ 160 |  |  |  |
| --- | --- | --- | --- | --- | --- | --- | --- | --- |
|  | Sensitivity | Specificity | PPV | NPV | Sensitivity | Specificity | PPV | NPV |
| **Abc19** | **86.6** | **61.5** | **90.3** | **52.5** | **95.7** | **42.3** | **63.8** | **90.2** |
|  | 81.3-90.2 | 47.0-72.9 | 85.5-93.5 | 39.3-63.5 | 90.8-98.1 | 33.7-49.9 | 56.8-69.3 | 79.8-95.6 |
| **Biomerica** | **89.2** | **53.4** | **88.0** | **56.4** | **95.8** | **35.5** | **60.4** | **89.1** |
|  | 84.3-92.4 | 39.9-64.7 | 83.0-91.4 | 42.3-67.8 | 91.0-98.1 | 27.6-42.8 | 53.7-65.9 | 77.8-95.1 |
| **Roche** | **98.2** | **27.6** | **83.7** | **80.0** | **100.0** | **14.7** | **54.9** | **100.0** |
|  | 95.4-99.4 | 16.7-38.8 | 78.6-87.3 | 56.3-92.9 | 97.4-100 | 9.2-20.6 | 48.6-60.1 | 83.2-100 |
| **LumiraDX** | **98.6** | **4.7** | **83.5** | **40.0** | **100.0** | **4.3** | **55.2** | **100.0** |
|  | 95.9-99.6 | 0.6-13.9 | 78.2-87.2 | 5.3-81.1 | 97.3-100 | 1.4-8.8 | 48.8-60.6 | 47.8-100 |

*Sensitivity, specificity, PPV and NPV for each LFA to predict NT<sub>50</sub> of ≥ 50 or ≥ 160. 95%* *confidence intervals (Clopper-Pearson) are also shown.*

The sensitivity of all the POCT for detecting an NT_50_ ≥ 50 (of the 280 serum samples, 222 had an NT_50_ ≥ 50 and 58 had an NT_50_ < 50) was at least 90%, and for Roche and LumiraDX it was > 98%. However, the NPV varied between 40 and 80%, as a result of the small number of samples that were negative by the POCT, relative to the number of positive samples (**Fig 4)**. Specificity performance was also poor with a large number of false positive results (POCT positive, but NT_50_ < 50). In particular, the LumiraDX assay had a specificity of < 5%, whereas the Abc19 assay had the highest specificity of 62%. These results impacted the PPV in this high prevalence setting, which ranges from 90% for the Abc19 assay and 84% for the LumiraDX assay.

When the NT_50_ threshold was raised to ≥ 160 (where 142 of the serum samples had an NT_50_ ≥ 160 and 138 had an NT_50_ < 160), the sensitivity for all the POCT rose to at least 96%, with a consequential increase in NPV to at least 90%. However, the specificity of all of the POCT fell, with the Abc19 kit having the highest specificity at 42%. This affected the PPV, which also fell from the lower NT_50_ threshold, to between 55 and 64%.

### Comparison of POCT device performance in serum and capillary samples

A major advantage of the POCT devices over immunoassays performed within accredited laboratories is the lack of requirement for phlebotomy. However, many evaluations of POCT performance have not compared serum and capillary samples. Furthermore, it has been suggested that LFA could be suitable for home use, outside of a healthcare setting, and this would require the participant to interpret their test result.

In this work, paired serum and capillary samples were available for a number of participants in the convalescent cohort, and the performance of seven POCT assays (Abc19, Biomerica, Biozek, Fortress, Menarini, Roche and LumiraDX) was compared. The serum sample was processed in the laboratory, and read by a health care worker, and the capillary sample was read by the study participant (with the aid of a labelled diagram or instruction leaflet) and a second health care worker in the research clinic. Sensitivity findings for the 3 matched interpretations are shown in **Table 5 and Fig 5**.

**Table 5.** Comparison of paired serum and capillary results on selected POC devices.

|  | ABC 19 | Roche | Menarini | Fortress | Biomerica | Biozek |
| --- | --- | --- | --- | --- | --- | --- |
| Matched capillary (participant) capillary (HCW) and serum (laboratory) results | N= 45 | N= 42 | N=38 | N=41 | N=11 | N=41 |
| Sensitivity (%) (95% CI) |  |  |  |  |  |  |
| Capillary (participant) | **33.3**<br>(20.0-49.0) | **64.3**<br>(48.0-78.4) | **63.2**<br>(46.0-78.2) | **61.0**<br>(44.5-75.8) | **18.2**<br>(2.3-51.8) | **39.0**<br>(24.2-55.5) |
| Capillary (HCW) | **53.3**<br>(37.9-68.3) | **69.0**<br>(52.9-82.4) | **71.1**<br>(54.1-84.6) | **58.5**<br>(42.1-73.7) | **9.1**<br>(0.2-41.3) | **46.3**<br>(30.7-62.6) |
| Serum (laboratory) | **73.3**<br>(58.1-85.4) | **88.1**<br>(74.4-96.0) | **76.3**<br>(59.8-88.6) | **46.3**<br>(30.7-62.6) | **36.4**<br>(10.9-69.2) | **73.2**<br>(57.1-85.8) |
| Kappa |  |  |  |  |  |  |
| Serum Vs Capillary (HCW) | **0.31**<br>(0.06-0.57)<br>p= 0.35 | **0.46**<br>(0.18-0.74)<br>p=0.008 | **0.59**<br>(0.30-0.88)<br>p=0.697 | **0.47**<br>(0.21-0.73)<br>p=0.227 | **-0.17**<br>(-0.46-0.12)<br>p=0.38 | **0.29**<br>(0.05-0.54)<br>p=0.007 |
| Serum Vs Capillary (participant) | **0.21**<br>(0.02- 0.40)<br>P<0.001 | **0.39**<br>(0.13-0.65)<br>p=0.002 | **0.57**<br>(0.30-0.84)<br>p=0.125 | **0.42**<br>(0.16-0.68)<br>p=0.146 | **0.12**<br>(-0.43-0.67)<br>p=0.63 | **0.38**<br>(0.17-0.59)<br>P<0.001 |
| Capillary (HCW) Vs Capillary (participant) | **0.52**<br>(0.29- 0.75)<br>p= 0.12 | **0.89**<br>(0.75-1.03)<br>p=0.50 | **0.82**<br>(0.63-1.01)<br>p=0.250 | **0.95**<br>(0.85-1.05)<br>p=1.000 | **0.62**<br>(-0.04-1.28)<br>p=1.000 | **0.85**<br>(0.69-1.01)<br>p=0.25 |

*Sensitivity for LFA devices comparing serum, and HCW and participant interpreted capillary* *data. Sensitivity and 95% CI (Clopper-Pearson). P values calculated using McNemar's test* *comparing serum and capillary sensitivity. Kappa describing extent of agreement between* *results.*

**Fig 5:**
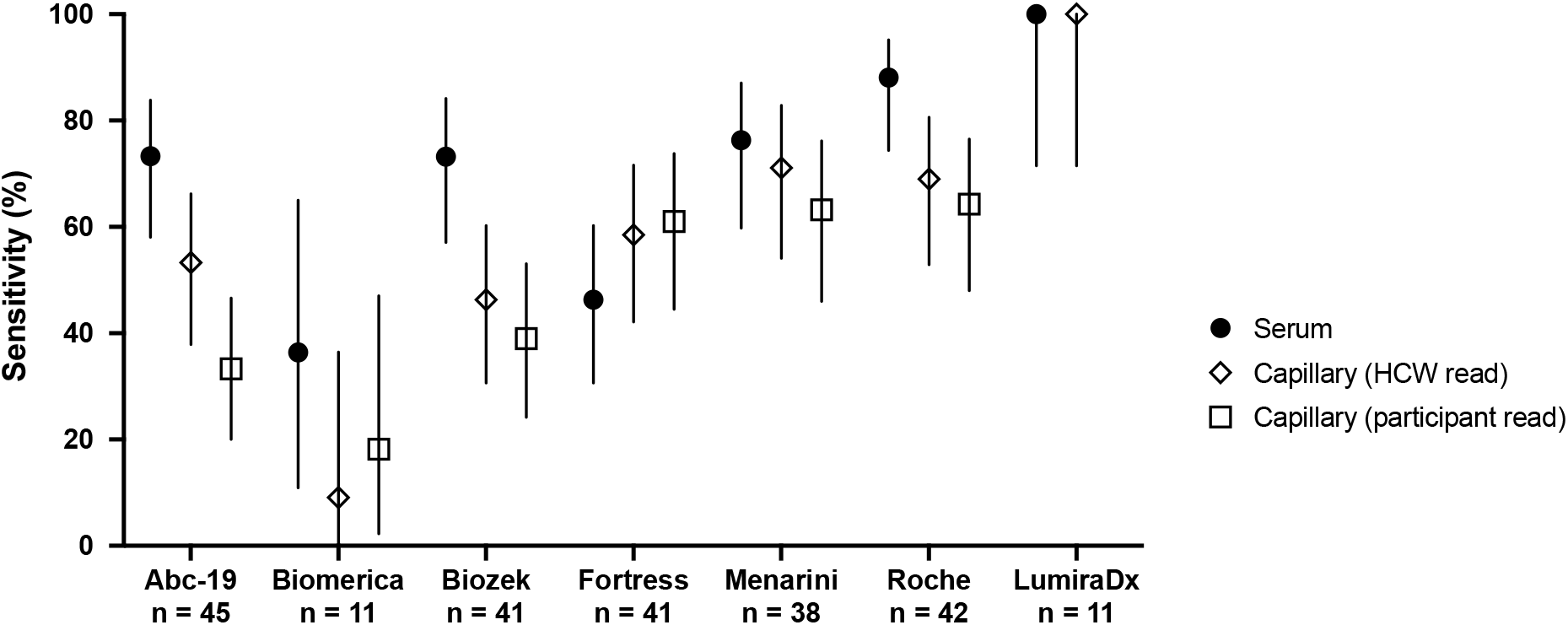
Sensitivity comparison between paired serum and capillary samples. Sensitivity for LFA devices comparing serum, and HCW and participant interpreted capillary data. Sensitivity and 95% CI (Clopper-Pearson) were plotted.

A common feature of the LFA results in capillary samples was a reduction in sensitivity compared to the serum samples. This was seen in five out of the 7 LFA analysed, however, the magnitude of the effect varied. Both Biozek and Roche showed significant decreases between serum and capillary sensitivity when interpreted by either HCW or participant. This decrease in sensitivity ranged from 19.1 to 34.2%. ABC 19 showed a significant decrease in sensitivity between serum and participant read capillary results decreasing from 73.3% to 33.3%. The Fortress LFA showed a small increase in sensitivity in capillary samples compared to serum but this was not statistically significant. The LumiraDX assay showed 100% sensitivity for both samples (n=11 but a large number of test fails (n=18) occurred on capillary samples at the research clinic.

The concordance between serum, HCW and participant read capillary results are displayed in table 5. Concordance between LFA varied. For example of the 45 Abc19 results, 24 were interpreted as positive by a HCW and 15 by the participant. In contrast, for the Fortress LFA, there was a single discrepant positive result between the HCW and participant interpretation, where the participant reported a positive, and the HCW a negative, result.

Participants who performed self-testing using the AbC-19 LFA in the clinic also completed a questionnaire that addressed aspects of self testing using the kits (n = 35 participants). Their responses are summarised in **Fig 6**. Overall, 89% of participants reported it was “very easy” or “easy’ to understand the leaflet explaining the results, and 75% reported it was “very easy” or “easy” to understand the instructions. Any issues appeared to be predominantly associated with taking the capillary samples and using the test kit, with 20% of participants reporting it was “difficult” or very difficult’ to take the sample.

**Fig 6:**
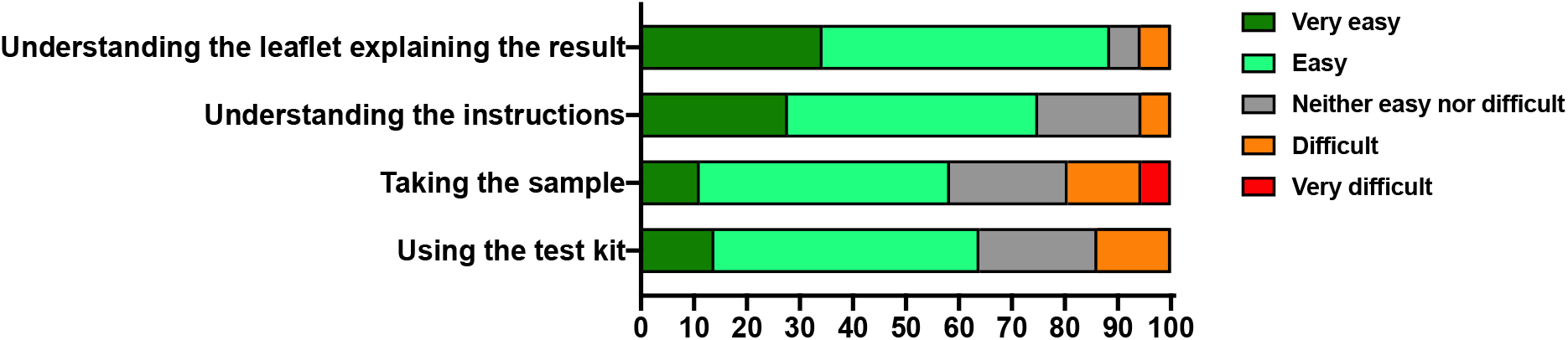
Ease of use. Summary of ease of use questionnaire. Participant ease of use responses to the indicated categories were expressed as a percentage of responses.

## Discussion

The SARS-CoV-2 pandemic has generated unprecedented demand for testing both to confirm acute infection and past virus exposure. In particular, serological assays measure prior exposure to the virus and are being used in high volume settings to measure seroprevalence at a population level. The role for these tests at an individual level is still poorly understood, and there are huge risks of using poorly performing tests. For example, using a test with a poor specificity in a low prevalence population would mean a high ratio of people with a false positive result compared to a true positive result, leading people to believe they were antibody positive, when they are not.

This study was designed as part of a national evaluation of SARS-CoV-2 antibody POCTs within NHS Scotland, and aimed to address several questions regarding the performance of the POCT. To be effective, POCT should have similar performance in samples from patients who experience relatively mild illness compared to patients requiring hospitalisation. This was addressed using a cohort of convalescent patients, the majority of whom did not require hospital treatment during their illness. This cohort was also used to assess the performance of the POCT with increasing time from infection, and to examine how well they were able to predict neutralising antibody titres.

Another major requirement is for POCT to perform well in capillary samples compared to serum. This point is critical since a major strength of the POCT devices is that they could be used on capillary samples, thus reducing the requirement for phlebotomy.

In this work, 14 POCTs were evaluated on serum samples, and a number of these tests underwent further evaluation on capillary samples. From the serum evaluation, it became clear that specificity performance of many of the POCT was good – with eleven out of fourteen reaching the MHRA specificity target of > 98% for IgG, or in the case of LumiraDX total Ab. However, only a single device (LumiraDX) reached the MHRA sensitivity criteria of >98% ≥ 20 days post symptom onset. The sensitivity panel that was used to assess these POCT in the first instance consisted predominantly of hospitalised patients, who tend to have higher antibody titres than patients with milder disease (17–22), making disease severity a less likely explanation for these observations. The reduced sensitivity of the LFA compared to manufacturer’s claims is not unique to the LFA; the sensitivity of a laboratory analyser immunoassay on this cohort was 93% at ≥ 20 days post symptom onset. Therefore at relatively early time points post symptom onset, a negative result cannot rule out SARS-CoV-2 exposure. Indeed, for the subgroup of LFA where sensitivity with time was examined it was clear that, at early time points, sensitivity increased with time reaching maximal sensitivity after 21 days post symptom onset. This was in keeping with other reports that have studied the time to reach maximal antibody titres (17,23–25).

An equally important question regarding timing of testing was how long post infection the POCT could detect an antibody response. The issue of sero-reversion is important for seroprevalence studies, and the ideal test for this purpose would have a high sensitivity for a long period of time. Otherwise, sero-reversion may result in underestimation of seroprevalence, and models to account for this trend of waning antibodies themselves risk over-or under-correcting for this effect (26,27). To examine this, a cohort of SARS-CoV-2 convalescent individuals for whom longitudinal serum samples were available were used. Crucially, these patients were most representative of the majority of SARS-CoV-2 infections in so far as only 5 participants providing serial samples required hospitalisation, and none required treatment in intensive care. In this work, the LumiraDX assay was the only assay to maintain a high sensitivity at time points from 21 to 244 days post symptom onset; the other assays showed variable levels of declining sensitivity. This was most marked for the Biomerica LFA, which had a sensitivity of 91% at 21-79 days post symptom onset, dropping to 54% at ≥ 140 days post symptom onset.

The relative importance of specificity compared to sensitivity at least in part depends on the intended use. For example, for serosurveillance, particularly in low prevalence populations, specificity should be maximised to reduce the false positive rate. However, in clinical settings, drops in specificity with increased sensitivity might be acceptable if there were adequate follow up tests to identify true positives.

A major question remains over what a positive antibody test result means in terms of protection from reinfection, or protection from infection following vaccination. This is pertinent given the continued discussions regarding the issue of “immunity passports” and a possible role for antibody tests in assisting populations where vaccine supply is scarce through identifying antibody negative individuals who should be prioritise for vaccination and/or antibody positive individuals who may be suitable for reduced vaccine dosing regimens. In this work, we compared the LFA result with the NT_50_ level (**Fig 4** and **Table 4**). It was apparent that a positive LFA was associated with a wide range of neutralising antibody levels, and that a number of LFA positive samples had low NT_50_ levels (< 50). This adds to the concern that a positive LFA result may not inform about the likelihood of protection from infection. Alongside specificity concerns and the increased circulation of variants of concern that may be able to evade existing humoral responses (28,29), this highlights the risk of using these tests for the purpose of immunity passports. Whilst some of the better performing LFA may be suitable for sero-prevalence studies, they should not be used to draw conclusions on an individual’s protection from reinfection.

A major advantage of POCT stems from their potential to relieve requirement for phlebotomy and to be performed outside of a healthcare setting. However, for this potential to be realised, additional factors including POCT performance in capillary samples and ease of use should be considered. In this work, a head to head comparison of POCT performance on serum and capillary samples was performed, and for the LFAs HCW and participant interpretation of the test result was also compared. There were differences in performance on serum and capillary samples, with the majority of LFA showing reduced sensitivity on capillary samples compared to serum. Concordance between serum and capillary results was sometimes poor indicating that that the performance of a test on serum under laboratory conditions cannot be assumed to equate to performance on a capillary sample. Only the LumiraDX device was equally sensitive on both capillary and serum samples.

There was good agreement between capillary results interpreted by HCW and participants for the Roche, Menarini, Biozek and Fortress kits studied indicating that these LFA tests may be best suited for use as home test kits as interpretation at home is likely to match interpretation in a hospital setting. For other LFAs, such as AbC-19, many participants had difficulty noticing the faint lines produced by the test kit and interpreted the result as being negative when it was interpreted as positive by a HCW.

A disadvantage of the POCT devices compared to laboratory immunoassays is that they are strictly qualitative (with the exception of LumiraDX where a numeric value is converted to a qualitative result), meaning the possibility of introducing equivocal zones is not available. Further issues that came to light during the course of this evaluation work included the possibility of batch-to-batch variation in kits from the same manufacturer. For example two different batches of Menarini kits showed marked differences in sensitivity (76.7% on one batch verus 94.9% on another batch). Whilst we did not observe any failed tests during our evaluation of the LFA using serum or capillary samples, a high failure rate was seen using the LumiraDX device with capillary samples with 18 out of 29 participants having a failed test result with lumiraDX. Unpublished data available from another POCT evaluation site in Scotland (NHS Tayside) where a newer version of the LumiraDX device was being trialled indicated that there were no issues with failed SARS-CoV-2 antibody capillary tests with the newer version of the LumiraDX device suggesting that this issue has now been resolved by the company. However, strategies to identify such issues should continue to be employed by the manufacturer and/or the end user to ensure accurate performance on capillary samples.

The strength of this work lies in its contribution to our understanding of POCT performance – in particular performance at different times post infection, confirmed evidence of batch to batch variation with some POCT kits and through demonstrating the general lack of correlation between a positive POCT result and the presence of neutralising antibodies. Our data also demonstrates that the results obtained in the lab using serum cannot automatically be assumed to be representative of finger-prick capillary test results. Furthermore, a good proportion of this work used a convalescent cohort of patients with relatively mild disease, making the findings applicable to community based studies. However, the study does have limitations. Due to sample availability constraints and the large number of POCTs evaluated not all could be evaluated on the same panel of positive and negative samples. In particular, the numbers of samples where paired serum and capillary sample results were available to assess concordance was limited for the majority of the POCT examined. For one of the test kits ease of use was assessed but as many of the study participants providing capillary samples in this study were healthcare workers the data obtained through this may not be representative of the general population.

In summary, our results highlight a wide variation in performance of SARS-CoV-2 antibody test kits and illustrates the importance of evaluating multiple different aspects of test performance including checking for batch to batch variation, changes in sensitivity as time from infection increases, correlation with neutralising antibodies and performance on capillary samples. Thorough evaluation of all these aspects is essential prior to considering the utilisation of these tests for antibody or ‘immunity’ passports and identification of hospitalised patients who would benefit from monoclonal antibody treatment.

## Data Availability

All data produced in the present study are available upon reasonable request to the authors

## Acknowledgements

Menarini test kits (batch batch 2003288) were supplied by Annette Thomas from Cardiff & Vale University Health board. Funding and support for this study was provided by Lothian NRS BioResource. We would also like to thank members of the National Service Scotland team for their support with co-ordinating this work.

## Abbreviations

LFA: (lateral flow assay)
POC: (point of care)
PPV: (positive predictive value)
NPV: (negative predictive value)
RT-PCR: (real time polymerase chain reaction)
NT50: (half maximal neutralising titre)
hCoV: (Human Coronavirus)
SARS-CoV-2: (severe acute respiratory syndrome coronavirus 2)
COVID-19: (coronavirus disease 2019).

## Supplementary information

S1 Table. **Manufacturers published product specifications and performance data**

