## Supporting table 1 for "Evaluation of SARS-CoV-2 Antibody Point of Care Devices in the Laboratory and Clinical Setting"

| Company | Product specifications | Manufacturer's Sensitivity | Manufacturer's Specificity |
|---|---|---|---|
| Abingdon Health Laboratories<br>2007003<br>Antibody type: IgG<br>Target antigen: Spike (S) protein | Transfer 2.5µL of venous whole blood or serum or 5µL capillary blood to sample well. Add 100µL buffer solution. Read results at 20 minutes. | 98.03% (95% CI: 95.03%-99.46%)<br><br>N= 203 positive cases on commercial IgG SARS-COV-2 ELISA kit | 99.56% (95% CI: 98.4%-99.95%)<br><br>N= 450 pre-pandemic samples |
| Alpha Pharma<br>VID35 08 012 LOT E2004009<br>Antibody type: IgG and IgM<br>Target antigen: S protein | Transfer 10µL sample (venous or capillary whole blood or serum) to sample well. Add 2 drops (60-80µL) buffer. Read result at 15 minutes- don't read after 20 minutes. | Infection time 4-10 days: 81.25%<br>Infection time 11-24 days: 97.1 %<br>N=397 PCR positive cases | 100%<br>N= 128 PCR negative samples |
| Biomerica<br>kit 1507A-50, BATCH 6630<br>Antibody type: IgG and IgM<br>Target antigen: Nucleocaspid (N) protein | Transfer 10µL serum or 20µL venous or capillary whole blood to sample well. Add 2 drops (80µL) buffer. Read at 10 minutes. Do not interpret results after 20 minutes. | 85.0% (95%CI: 62.1%-96.8%)<br>N= 20 PCR positive cases | 96.3% (95%CI: 89.4%-99.2%)<br>N= 80 PCR negative cases |
| Biozek<br>BNCP402E00084<br>Antibody type: IgG/IgM<br>Target antigen: not disclosed | Transfer 20µL whole blood or 10µL serum/plasma to sample well. Apply 2 drops (80µL) buffer. Read results at 10 minutes. Do not interpret after 20 minutes. | IgG 100% (95% CI: 86.0-100%)<br>IgM 85.0% (95%CI: 62.1-96.8%)<br>N=20 PCR positive cases | IgG 98.0% (95%CI: 89.4%-99.9%)<br>IgM 96.0% (95%CI: 86.3-99.5%)<br>N=50 negative PCR cases |
| Fortress<br>COV19-OTC-2004168-1<br>Antibody type: IgG and IgM<br>Target antigen: S protein | Transfer 5-10µL whole blood, serum or plasma into the sample well. Add 2 drops of buffer and read results at 10 minutes. | 95.6% (90.7- 98.4%)<br>N=137 | 95.2% (91.5-97.7%)<br>N= 209 |
| Jiangsu<br>28d2001<br>Antibody type: IgG/IgM<br>Target Antigen: S protein | Transfer 20µL blood into sample diluent. Shake well then add 5 drops (100µL) of diluent/sample mixture to sample well. Read result at 3 minutes. | 97.14% (95% CI: 90.17%~99.21%)<br>N=70 positive samples | 100% (95% CI: 93.98%~100.00%)<br>N=60 negative samples |
| Lepu<br>20CG2508x<br>Antibody type: IgG/IgM<br>Target Antigen: not disclosed | Transfer 10µL serum or plasma or 20µL whole blood to sample well. Add 2 drops (80µL) buffer. Read test result at 10-20 minutes. Result should not be interpreted after 20 minutes. | IgG sensitivity: 100%<br>N=92 Positive samples with reference LFIA method<br>IgM sensitivity: 97%<br>N= 72 positive samples reference LFIA method | IgG specificity: 99%<br>N=128 negative samples with reference LFIA method<br>IgM specificity 100%<br>N=147 negative samples with reference LFIA method |
| Menarini / Healgen<br>GCCOV402A (2005156)<br>Antibody type: IgG/IgM<br>Target antigen: S1, S2 and N protein | Transfer 5µL serum/plasma or 10µL whole blood to sample well. Add 2 drops (80µL) buffer to buffer well. Read at 10 minutes and no later than 15 minutes. | IgM sensitivity 87.9% (87/99)<br>N= 99 PCR positive samples<br>IgG sensitivity 97.2% (35/36) during the convalescence period | IgG/IgM specificity is 100%( 14/14)<br>N=14 PCR negative samples |
| Menarini 2<br>GCCOV402A (2003288)<br>Antibody type: IgG/IgM<br>Antigen target: S1, S2 and N protein | Transfer 5µL serum/plasma or 10µL whole blood to sample well. Add 2 drops (80µL) buffer to buffer well. Read at 10 minutes and no later than 15 minutes. | IgM sensitivity 87.9% (87/99)<br>N= 99 PCR positive samples<br>IgG sensitivity 97.2% (35/36) during the convalescence period | IgG/IgM specificity is 100%( 14/14)<br>N=14 PCR negative samples |
| Mologic/ Visitec<br>7066351<br>Antibody type: IgG/IgM/IgA<br>Antigen target: S2, RBD (receptor binding domain), N protein | Transfer 5µL whole blood/plasma/serum to sample well. Add 2 drops of buffer. Read results at 10 minutes. | 96% (79.65-99.9%)<br>N=25 PCR confirmed cases | 98.8% (99.6-99.8%)<br>N=257 pre-COVID 19 pandemic samples |
| Roche<br>9901-ncov-02c<br>qc07920001<br>Antibody type: IgG/IgM<br>Target Antigen: S protein, N protein | Transfer 20µL whole blood or 10µL serum/plasma to sample well. Apply 3 drops (90µL) buffer. Read results between 10 and 15 minutes. | 7-14 days 92.59% (82.11-97.94%)<br>N= 54 PCR positive samples<br>>14 day sensitivity 99.03% (94.71-99.98%)<br>N=103 PCR positive samples | 98.65% (96.1-99.72%)<br>N=222 PCR negative samples |
| Pharmact<br>1797 6618<br>Anti-body type: IgG/IgM<br>Target antigen: not disclosed | Transfer 50µL whole blood or serum to sample well. Add 2 drops of buffer. Read at 20 minutes. | 4-10 days symptom onset: 70% for IgM<br>11-24 days symptom onset: 92.3% for IgM<br>11-24 days symptom onset: 98.6% for IgG | 100%<br>N=126 |
| idsolid<br>No batch numbers on kits<br>Antibody type: IgG/IgM<br>Antigen target: not disclosed | Transfer 20µL whole blood or 10µL serum/plamsa to sample well. Apply 2 to 3 drops of buffer (about 100µL). Read results at 15 minutes. | IgM sensitivity 96% (48/50)<br>IgG sensitivity 98.2% (56/57)<br>reference test PCR | IgM specificity 100% (100/100)<br>IgG specificity 100% (100/100) |
| Wuhan Life Origin Biotech/Szybio<br>C200525001/SF20025<br>Antibody type: IgG/IgM<br>Target Antigen: not disclosed | Transfer 20µL whole blood or 10µL serum/plamsa to sample well. Apply 2 drops (60µL) buffer. Read result at 15 minutes and no later than 18 minutes. | not reported | not reported |
| Wuhan Easy diagnostics<br>SA-2-D 20050602<br>Antibody type: IgG/IgM<br>Target Antigen: not disclosed | Transfer 10µL plamsa/serum or 15µL whoel blood to sample well. Add 2 drops (70µL) sample diluent. Read result at 10 minutes. | 100% (95% CI: 100-100%)<br>N=207 PCR positive samples | 99.56% (95% CI: 98.94-100%)<br>N=229 PCR negative samples |
